# Age and Sex Differences in the Genetics of Cardiomyopathy

**DOI:** 10.1101/2021.04.06.21255002

**Authors:** Oyediran Akinrinade, Genomics England Research Consortium, Jane Lougheed, Tapas Mondal, John Smythe, Luis Altamirano-Diaz, Erwin Oechslin, Seema Mital

## Abstract

**Aims:** Cardiomyopathy is a clinically and genetically heterogeneous disorder with age and sex-related differences in severity and outcomes. The aim of our study was to identify age and sex-related differences in the genetic architecture of cardiomyopathy.

**Methods and Results:** We analyzed whole genome sequence data from 471 pediatric and 926 adult cardiomyopathy patients from our Heart Centre Biobank and from the Genomics England cohort. Overall yield of rare deleterious coding variants was higher in pediatric compared to adult onset cardiomyopathy, but not different by sex. *MYH7, TNNT2, MYL3*, and *VCL* variants were more frequent in pediatric patients; *TTN* and *OBSCN* variants were more frequent in adult patients, with *MYH7* (Odds ratio 3.6; CI 2.1-6.3) and *OBSCN* (Odds ratio 5.5, CI 2.0-21.4) remaining significant after adjusting for multiple testing. Variants in early-onset cardiomyopathy clustered in highly constrained coding regions compared to those in adult patients (p=3.9×10^−3^). There were also differences between pediatric and adult patients in variant location within *MYH7* and *TTN* genes. When analyzed by sex, variants in female compared to male patients were in more highly constrained coding regions (p=0.002).

**Conclusion:** Our findings highlight under-appreciated genetic differences in early versus late onset cardiomyopathy. Variants in childhood cardiomyopathy and in female patients were in highly constrained coding regions of the genome suggesting greater deleterious effects and strong purifying selection in the general population. Knowledge of the affected gene, variant location within the gene, and variant constraint scores may be useful in predicting early versus late onset cardiomyopathy.

## Introduction

Cardiomyopathies are genetic disorders of heart muscle that cause the heart to become dilated and weak, or hypertrophied and stiff. Phenotypically, there are five subtypes - dilated (DCM), hypertrophic (HCM), restrictive (RCM), left ventricular non-compaction cardiomyopathy (LVNC) and arrhythmogenic cardiomyopathy (ACM), with a significant overlap between phenotypes and genotypes.^1^ Together, cardiomyopathies are the most frequent indication for heart transplantation and the leading cause of sudden cardiac death.^2-6^ Variants in over 100 genes have been associated with cardiomyopathy. While cardiomyopathies can be secondary to infection, inflammation, syndromic disorders, neuromuscular disorders, inborn errors of metabolism, or drug toxicities, primary cardiomyopathies i.e. those isolated to the heart, are usually genetic, can be sporadic or familial, and usually have an autosomal dominant inheritance.^7-9^

Cardiomyopathies have high childhood penetrance with variable expressivity resulting in intra-familial and inter-familial variability in the age of onset and severity.^10, 11^ Studies have also reported sex-related differences with a higher incidence and severity of cardiomyopathy in males compared to females.^12-17^ While age-related penetrance, lifestyle factors, and hormonal differences can explain part of the clinical variability by age and sex, a systematic comparison of the genetics of childhood versus late onset cardiomyopathy or of disease in females versus males has not been performed.

Growth in publicly available large-scale human genomic variation datasets has enabled identification of disease-associated genes through case-control burden analyses, and provides an opportunity to study the distribution of variation within genes, identify mutational hotspots,^18, 19^ and define genes that are intolerant of variants through gene-level metrics such as Residual Variation Intolerance Score (RVIS) and pLI (probability of being loss of function intolerant) score.^19, 20^ Additionally, Havrilla *et al*. developed a map of constrained coding regions in the human genome that are under strong purifying selection and show a scarcity of variants in the general population.^21^ Variants found in these constrained regions are more likely to be associated with developmental and disease phenotypes.

The goal of our study was to evaluate age and sex-related differences in the genetics of cardiomyopathy by comparing the frequency and distribution of rare, deleterious variants within known cardiomyopathy genes and within constrained regions of these genes between pediatric and adult patients, and between male and female patients.

## Methods

### Study cohort

The study cohort comprised 1397 unrelated primary cardiomyopathy patients (471 were pediatric i.e. <18 years old at diagnosis; 926 were adult); 236 were consented through the multi-center SickKids Heart Centre Biobank Registry and 1161 patients were from the 100,000 Genomes Project accessed through the Genomics England Clinical Interpretation Partnership (version 8).^22, 23^ Cardiomyopathy phenotypic subtypes were defined by published clinical criteria.^3, 24^ Written informed consent was obtained from all biobank participants and/or their parents or legal guardians and the research protocol was approved by the Institutional Research Ethics Boards at the Hospital for Sick Children, Children’s Hospital of Eastern Ontario, Toronto General Hospital, London Health Sciences Centre, Kingston General Hospital, and Hamilton Health Sciences Centre. Patients with secondary cardiomyopathies resulting from inborn errors of metabolism, mitochondrial disorders, syndromic, and neuromuscular etiologies were excluded.

### Whole genome sequencing and variant interpretation

Whole genome sequencing (WGS) was performed on DNA derived from blood or saliva using Illumina HiSeq X to an average depth of 30X. WGS quality control, data processing, and variant identification were done as previously described.^25^ Identified variants were annotated using Ensembl’s Variant Effect Predictor (VEP v92).^26^ Rare missense variants predicted deleterious or damaging by SIFT and PolyPhen tools with Combined Annotation Dependent Depletion score of at least 20 were classified as “deleterious”.^27-29^ Variants predicted to cause loss-of-function (LoF) i.e. frameshift, nonsense, and splice-site variants, were annotated by LOFTEE via VEP, and rare variants flagged as high confidence LoF by LOFTEE were classified as deleterious.^30^ Rare variants were defined based on Minor Allele Frequency (MAF) <0.01% in Exome Aggregation Consortium reference individuals. Only coding single nucleotide variants (SNVs) and small insertion-deletions were assessed in this study.

### Cardiomyopathy candidate gene sets

Using information from Online Mendelian Inheritance in Man database, published literature, and genes represented on commercially available cardiomyopathy gene panels, we manually curated a list of 133 cardiomyopathy candidate genes (**Supplementary Table S1**). 56 genes with strongest association with cardiomyopathy were classified as Tier 1; these included the sarcomeric genes. Based on gene ontology, we grouped the 133 genes into 14 functional categories.

### Variants mapping to protein domains and constrained coding regions

Variants identified in genes with multiple transcripts were reported with respect to the canonical transcripts, and were mapped to uniprot protein domains. Protein domain annotation for all canonical transcripts of cardiomyopathy genes were obtained by mining the uniprot database (http://www.uniprot.org) (assessed May 28, 2020) using a custom in house script.^31^ We accessed the constrained coding region (CCR) map as published by Havrilla et al.^21^, and using this map, we obtained CCR score for all variants. By converting genomic coordinates to protein coordinates, we used the CCR map to obtain a measure of constraint on protein domains of cardiomyopathy genes.

### Statistical analysis

All rare coding SNVs and insertions-deletions predicted deleterious or high risk for altering protein function of known cardiomyopathy genes were used in the final analysis.Variant yield and burden of multiple variants in pediatric versus adult, and male versus female cases were compared for all cardiomyopathy patients as well as for the subset with only HCM and DCM. CCR scores were compared using Kolmogorov-Smirnov (KS) two-tailed test to determine if variants from different subgroups had variable distribution in cardiomyopathy patients and in reference control genomes available through the Genome Aggregation Database (gnomAD v2.1) (n=125,748). The uniformity of the spatial distribution of deleterious variants within proteins was assessed by KS Goodness-of-Fit Test. Comparisons between groups were performed with either χ^2^ test or Fisher’s exact test. Continuity correction was applied via Visualizing Categorical Data (vcd) R package (v1.4-8) to estimate odds ratio and confidence interval for any zero cells in the contingency table.^32^ Benjamini–Hochberg false discovery rate correction was performed on all analyses to adjust for multiple testing. All statistical analyses were performed using R statistical software (version 3.5.2).

## Results

### Study population

Whole genome sequencing data was available on 1397 eligible cardiomyopathy patients. 471 (34%) were pediatric cases, and 871 (62%) were male. By subtype, 768 (55%) had HCM, 435 (31%) had DCM, 116 (8%) had ACM, 58 (4%) had LVNC, and 20 (1%) had RCM. The cohort characteristics are described in **Table 1**.

**Table 1.** Characteristics of study cohort.

|  | **All (n=1397)** | **Pediatric**<br>**(n=471)** | **Adult**<br>**(n=926)** | **p-value** |
| --- | --- | --- | --- | --- |
| Mean age at diagnosis (years) | 31.4 ± 21.8 | 4.8 ± 5.9 | 44.0 ± 13.7 | 2.2 x 10<sup>-16</sup> |
| Male, n (%) | 871 (62.3) | 284 (60.3) | 587 (63.4) | 0.28 |
| Cardiomyopathy subtypes, n (%) |  |  |  |  |
| HCM | 768 (55.0) | 223 (47.3) | 545 (58.9) | 5.6 x 10<sup>-5</sup> |
| DCM | 435 (31.1) | 175 (37.2) | 260 (28.1) | 6.7 x 10<sup>-4</sup> |
| ACM | 116 (8.3) | 20 (4.2) | 96 (10.4) | 1.4 x 10<sup>-4</sup> |
| LVNC | 58 (4.2) | 33 (7.0) | 25 (2.7) | 2.4 x 10<sup>-4</sup> |
| RCM | 20 (1.4) | 20 (4.2) | - |  |
| **n (%) of variant-positive\* patients in all genes** |  |  |  |  |
| All cardiomyopathy (n=1397) | 635 (45.5) | 218 (46.3) | 417 (45.0) | 0.70 |
| HCM (n=768) | 330 (43.0) | 105 (47.1) | 225 (41.3) | 0.16 |
| DCM (n=435) | 215 (49.4) | 79 (45.1) | 136 (52.3) | 0.17 |
| **n (%) of variant-positive\* patients in Tier 1 genes** |  |  |  |  |
| All cardiomyopathy (n=1397) | 395 (28.3) | 150 (31.8) | 245 (26.5) | 0.04 |
| HCM (n=768) | 201 (26.2) | 75 (33.6) | 126 (23.1) | 0.004 |
| DCM (n=435) | 144 (33.1) | 56 (32%) | 88 (34%) | 0.77 |

*\*Variant positive = presence of a rare, deleterious variant in a cardiomyopathy gene*
*p value for pediatric versus adult cases*
*HCM, hypertrophic cardiomyopathy; DCM, dilated cardiomyopathy; ACM, arrhythmogenic cardiomyopathy; LVNC, left ventricular non-compaction cardiomyopathy; RCM, restrictive cardiomyopathy*

### Genetic differences in early versus late onset cardiomyopathy

#### Higher frequency of deleterious variants in pediatric cardiomyopathy

Overall, 635 patients (45% of the cohort) harbored one or more rare deleterious variants across 111 genes. Of these, 395 (28% of the overall cohort) harbored variants in 52 of the 56 Tier 1 genes. A higher proportion of pediatric patients harbored a deleterious variant in a Tier 1 gene compared to adults (32% vs 27% respectively, p=0.04). When analyzed by cardiomyopathy subtypes, a higher proportion of pediatric HCM cases harbored a deleterious variant compared to adult HCM (34% vs 23% respectively, p=0.004); this difference was not seen in DCM cases (**Figure 1a**). There was no significant difference in the proportion of patients harboring multiple deleterious variants between pediatric and adult cases (**Figure 1b**).

**Figure 1.**
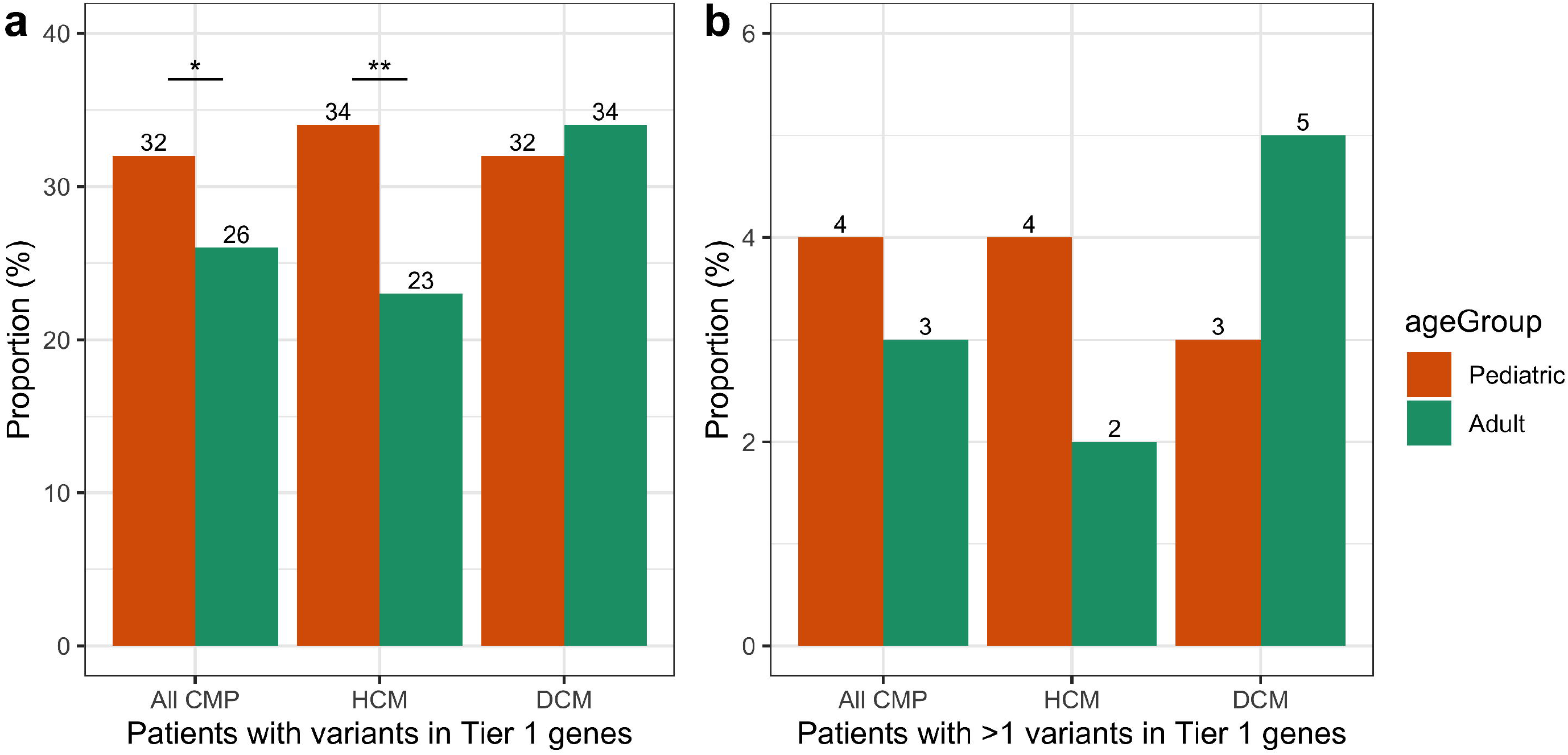
Variant yield by cardiomyopathy phenotypes. (**a**) Patients with variants in Tier 1 genes (471 pediatric, 921 adult cases). A higher proportion of pediatric patients (brown) harbored rare deleterious variants in Tier 1 cardiomyopathy genes compared to adult patients (green) (p=0.04). (**b**) There was no difference in frequency of multiple deleterious variants in cardiomyopathy genes between pediatric and adult cases. *p<0.05; **p<0.001 CMP, cardiomyopathy; HCM, hypertrophic cardiomyopathy; DCM, dilated cardiomyopathy

#### Higher variant CCR scores in pediatric cardiomyopathy

We obtained the CCR score for all deleterious variants identified in cardiomyopathy genes in our study cohort and in gnomAD. As expected, deleterious variants identified in cardiomyopathy patients were in more constrained coding regions compared to those in the gnomAD reference population i.e. had higher CCR scores (KS test p=2.2×10^−14^) (**Figure 2a, c**). This difference was also seen in the subset of Tier 1 genes (KS test p=1.1×10^−15^) (**Figure 2b, d**). For all genes and gene groups with at least 10 variants, we generated variant CCR score density maps (**Figures 3a-d**) and compared CCR score probability distribution using KS two-tailed test (**Figures 3e-h**). In the cardiomyopathy cohort, variant CCR scores across all cardiomyopathy genes was higher in pediatric compared to adult cases (KS test p=0.0039) and also when limited to variants in Tier 1 genes (KS test p=0.0005). CCR scores of variants in sarcomeric genes were also higher in pediatric versus adult cases (KS test p=0.0031). Variant CCR scores of non-sarcomeric genes did not differ by age (KS test p=0.27). Overall, these findings suggest that those with early onset disease are more likely to harbor variants within highly constrained coding regions.

**Figure 2.**
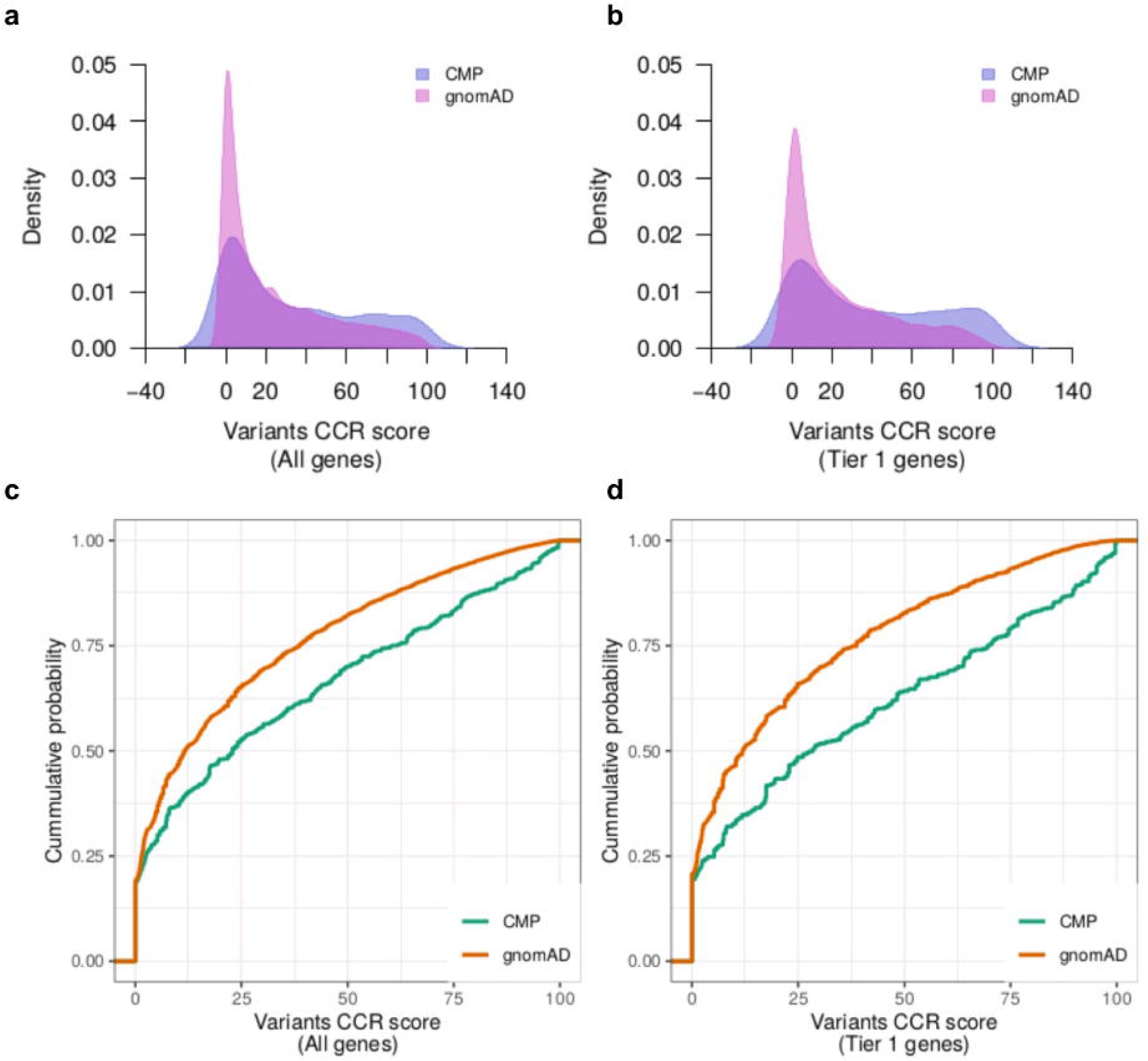
Constrained coding region (CCR) score of deleterious variants in all cases and controls. (**a, b**) Density plots of CCR scores, and (**c, d**) Empirical cumulative probability distribution of CCR scores of deleterious variants identified in cardiomyopathy cases (n=1397) and in gnomAD reference controls (n=125,748). Variant CCR scores were higher in cardiomyopathy cases compared to gnomAD reference controls across all cardiomyopathy genes (KS test p=2.19×10^−14^) and in the subset of Tier 1 genes (KS test p=1.11×10^−15^). CCR, constrained coding regions; CMP, cardiomyopathy; gnomAD, Genome Aggregation Database; KS, Kolmogorov–Smirnov test

**Figure 3.**
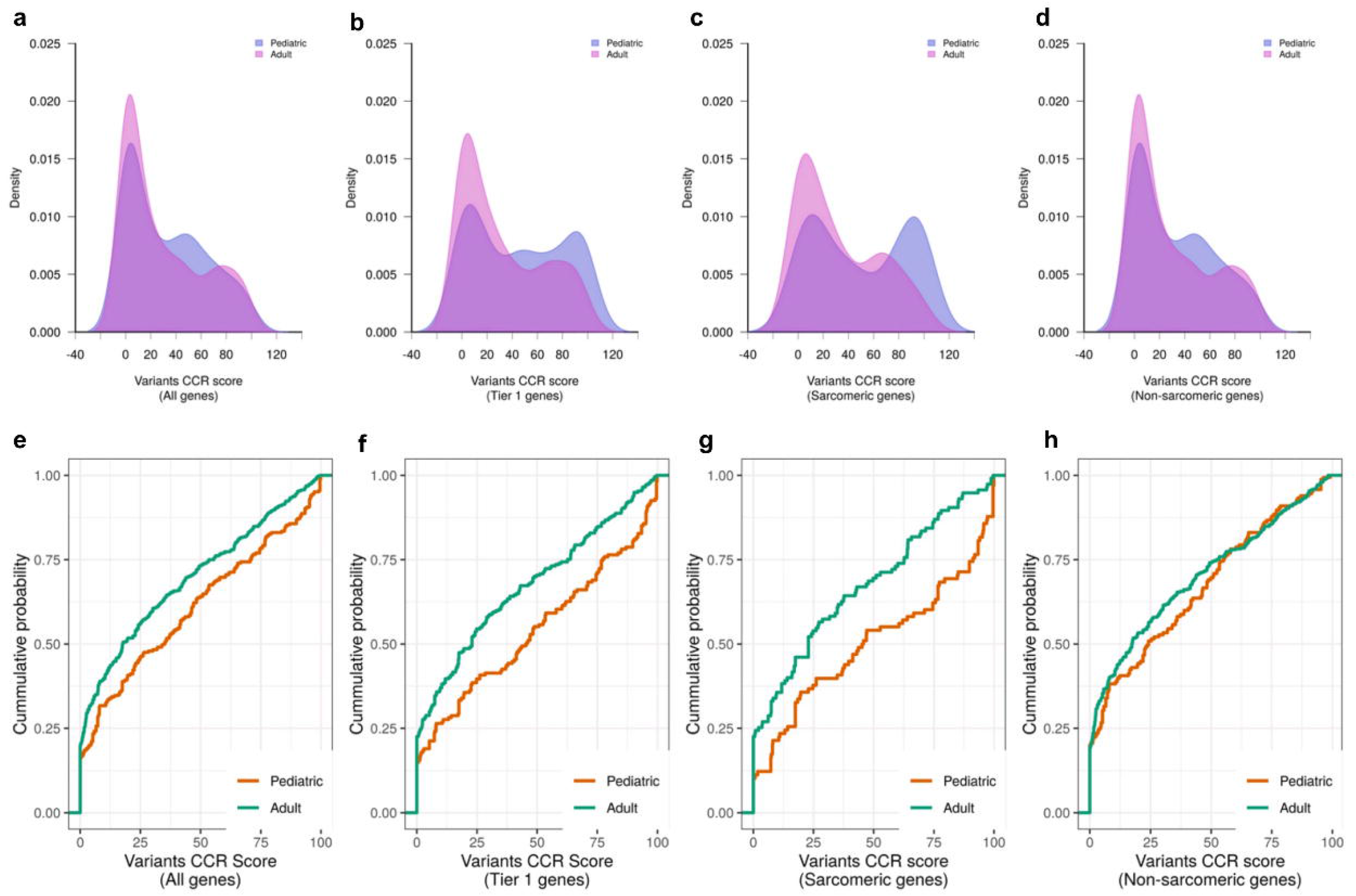
CCR score of deleterious variants stratified by age. (**a-d**) Density plots of CCR scores, and (**e-h**) Empirical cumulative probability distribution of CCR scores of deleterious variants identified in pediatric (n=471) and adult cardiomyopathy cases (n=921). Variant CCR scores were higher in pediatric versus adult cardiomyopathy cases across all cardiomyopathy genes (KS test p=0.0039), in the subset of Tier 1 genes (p=0.0005) and in sarcomeric genes (p=0.0031), but not different in non-sarcomeric genes (p=0.271). CCR, constrained coding regions; CMP, cardiomyopathy; KS, Kolmogorov–Smirnov test

#### Differentially affected genes in pediatric and adult cardiomyopathy

For gene level analysis, we compared the proportion of patients harboring at least one deleterious variant in a given gene. **Figures 4a-c** show the proportion of pediatric and adult patients harboring deleterious variants by gene and by variant types. **Figures 4d-f** show volcano plots by cardiomyopathy subtype for differentially mutated genes between pediatric and adult cases. Five genes were differentially mutated between pediatric and adult cases with *MYH7* being the most frequently mutated gene in overall pediatric cardiomyopathy cases. *TNNT2, MYL3*, and *VCL* were also more frequently mutated in pediatric versus adult patients (**Figure 4d, Supplementary Table S2**). Of note, LoF variants in *VCL* were only seen in children, not in adults (OR=13.8, p=0.038). In contrast, *TTN* truncating variants and *OBSCN* deleterious variants were more frequent in adults compared to children with cardiomyopathy (**Figure 4d, Supplementary Table S2**). Both *MYH7* and *OBSCN* variant burden differences remained significant after correction for multiple testing.

**Figure 4.**
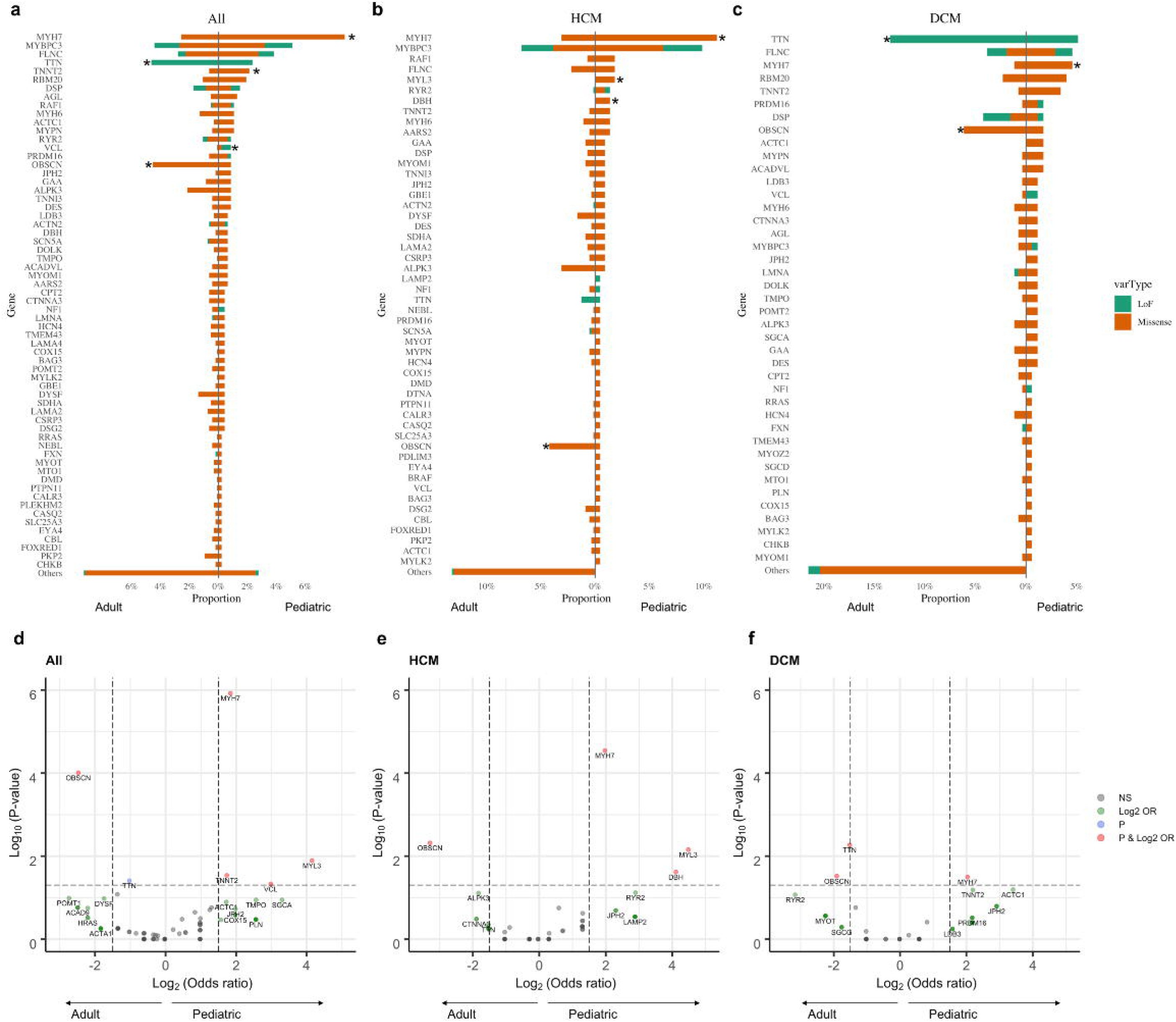
Genes harboring deleterious variants stratified by age. Bar plots showing genes harboring deleterious variants in patients with (**a**) any cardiomyopathy (n=1397), (**b**) only hypertrophic cardiomyopathy (n=768), and (**c**) only dilated cardiomyopathy (n=435). *p<0.05 between pediatric and adult patients. (**d-f**) Volcano plots showing log odds ratio of the frequency of variants between pediatric and adult patients with (**d**) any cardiomyopathy, (**e**) hypertrophic cardiomyopathy only, and (**f**) dilated cardiomyopathy only. *MYH7, TNNT2* and *VCL* were more frequently mutated in pediatric patients, and *TTN* and *OBSCN* were more frequently mutated in adult patients. NS, no significant difference (grey); log2 OR, genes meeting log2 (OR) ≥ 1.5 between pediatric and adult (green); P, genes with p<0.05 for variant burden between pediatric and adult (blue); p & log2 OR - genes with p<0.05 for variant burden and with at least 1.5 odds ratio between pediatric and adult cases (red). HCM, hypertrophic cardiomyopathy; DCM, dilated cardiomyopathy; OR, odds ratio

Subgroup analysis by cardiomyopathy subtype showed that *MYH7, MYL3* and *DBH* were more frequently mutated in pediatric HCM, while *OBSCN* was more frequently mutated in adult HCM (**Figure 4e, Supplementary Table S2**). *TTN* was the most affected gene in adult DCM accounting for 13.5% of cases compared to 5.1% of pediatric DCM. *OBSCN* was the second most affected gene in adult DCM with a higher variant burden compared to childhood DCM (**Figure 4f, Supplementary Table S2**). Of note, *FLNC*, a gene that is not routinely captured in all commercially available gene testing panels, was involved in 4.1% of all DCM cases. Overall, these findings suggest that variants in several sarcomeric and structural genes are more likely to be associated with early onset disease.

#### Variant burden by gene category in pediatric and adult cardiomyopathy

When analyzed by gene groups, sarcomeric genes were more frequently mutated in pediatric patients, particularly in the subset with HCM (**Figure 5, Supplementary Table S3**). Conversely, desmosomal genes were more frequently mutated in adult cardiomyopathy, particularly in the subset with DCM. Ion channel genes were also more frequently mutated in adult DCM compared to pediatric DCM (**Figure 5, Supplementary Table S3**).

**Figure 5.**
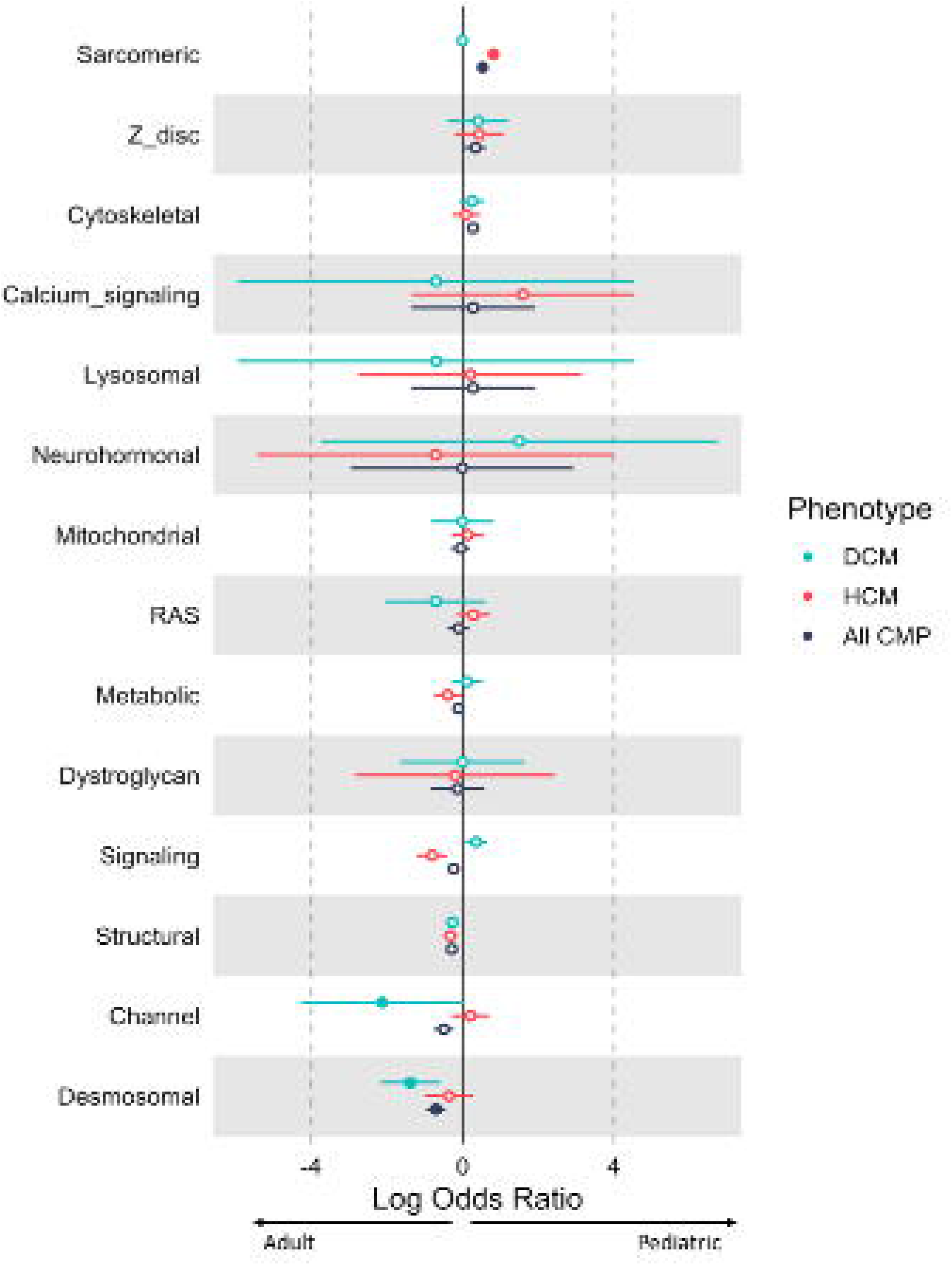
Gene categories harboring rare deleterious variants stratified by age. The forest plot shows the log odds ratio comparing proportion of variant carriers between 471 pediatric and 926 adult patients for different gene categories across different cardiomyopathy subtypes. Sarcomeric genes were more frequently mutated in pediatric cases; desmosomal genes were more frequently mutated in adult patients. Filled circles indicate significant differences between pediatric and adult patients. CMP, cardiomyopathy (black); DCM, dilated cardiomyopathy (blue), HCM, hypertrophic cardiomyopathy (red)

#### Variant location within protein domains

We used goodness of fit test to determine if deleterious variants were uniformly distributed within protein domains of genes or clustered in mutational hotspots within genes. This was analyzed only for genes with a high variant burden i.e. >10, in the overall cohort. **Figure 6** shows the spatial distribution of deleterious variants in *MYH7, MYBPC3, TTN*, and *OBSCN*. Deleterious variants were non-uniformly distributed in *MYH7* in pediatric patients with variants clustering within the myosin head and neck domains (KS test p=8.4×10^−4^). In adults, variants were more uniformly distributed across myosin head, neck and tail domains (KS test, p=0.058) (**Figure 6a**). For *MYBPC3*, variants clustered within the C5, C7, and C10 domains but this clustering was not different between children and adults (**Figure 6b**). *TTN* truncating variants were non-evenly distributed with clustering in the A-band domain (p=3.4×10^−4^) as has been previously described.^33-35^ Within the A-band, there was non-uniform variant distribution in adults (p=3.5×10^−3^) but not in children (p=0.49) (**Figure 6c**). *OBSCN* deleterious variants were non-evenly distributed and clustered within the protein kinase domain of the gene in adult patients (p=0.033) (**Figure 6d**). Overall, these findings suggest that location within protein domains may influence age-related penetrance of a variant.

**Figure 6.**
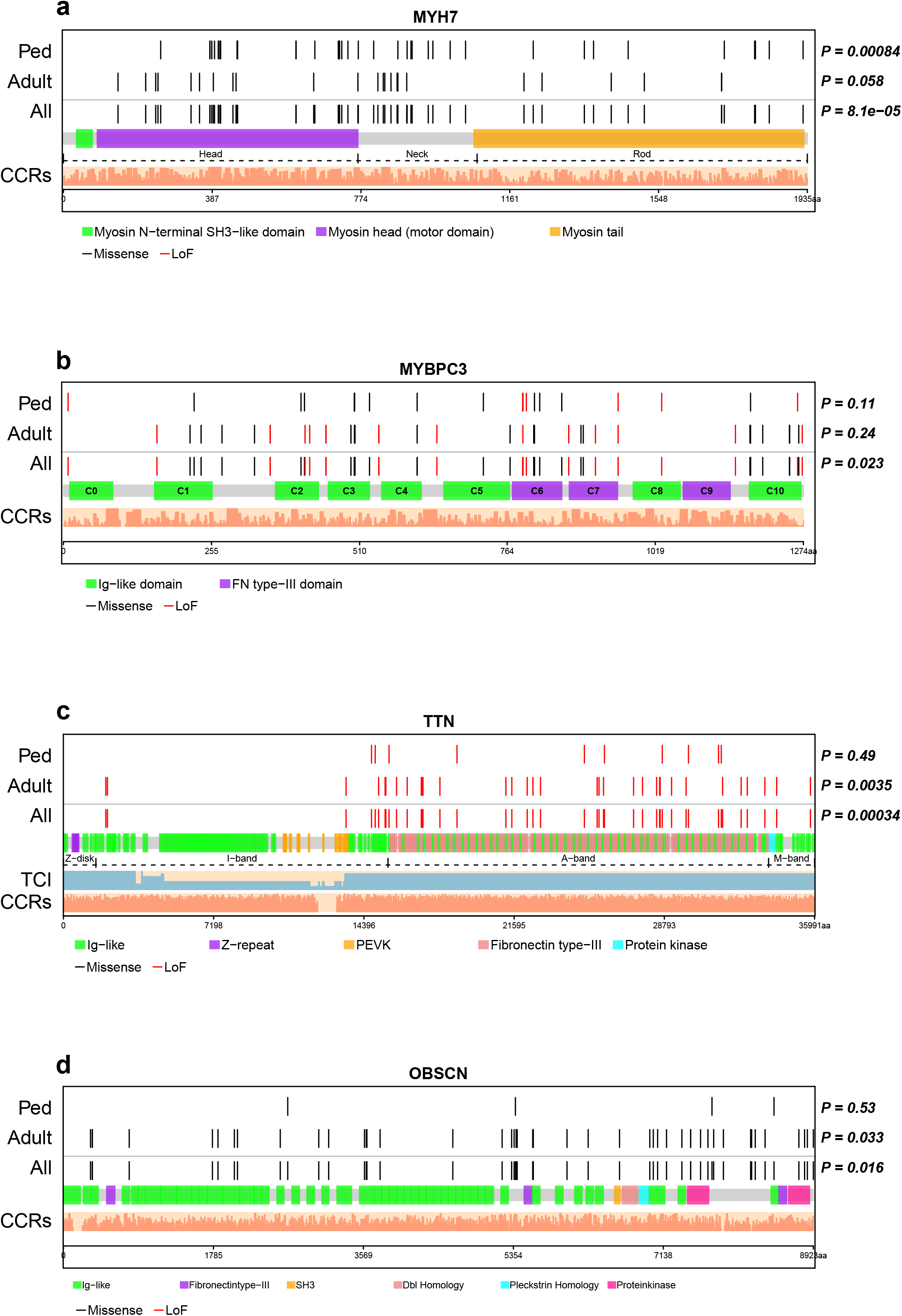
Spatial distribution of deleterious variants within protein domains. Each protein is linearly depicted with uniprot domain information. Light salmon bars show the constrained coding region scores across the entire protein. (**a**) MYH7 showing head, neck and rod/tail regions. Unlike variants in adult patients, deleterious variants in pediatric patients were non-uniformly distributed with clustering within head and neck domains and fewer variants in the tail domain (p=0.00084). (**b**) MYBPC3 showing the immunoglobulin and fibronectin type 3 domains. Deleterious variants clustered in C5, C7, and C10 domains (p=0.023), but this distribution did not differ between pediatric (n=471) and adult (n=926) patients. (**c**) TTN showing the four regions and all domains. The light blue bars under titin show the transcript count index. TTN truncating variants clustered in the A-band domain in adult patients (p=0.0035) but were uniformly distributed in pediatric patients. (**d**) OBSCN domains. Deleterious variants clustered within the protein kinase domains of the genes in adult patients (p=0.033) but not in pediatric patients. Non-random distribution of variants with each protein was assessed using Kolmogorov–Smirnov goodness-of-fit test. CCR, Constrained coding region; TCI, transcript count index

## Genetic differences by sex

There was no difference in overall variant yield between male and female patients (871 males, 526 females). There were no sex-related differences in variant CCR scores in gnomAD reference genomes (p=0.51) (**Figure 7a, d**). However, in the cardiomyopathy cohort, the deleterious variant CCR scores were higher in females compared to males. This was true of variants in all cardiomyopathy genes (p=2.5×10^−3^) (**Figure 7b, e**) as well as for variants in Tier 1 genes (p=0.019) (**Figure 7c, f**). Interestingly, we found that deleterious variants in LoF intolerant genes identified in female cardiomyopathy patients clustered in more constrained regions than those identified in male cardiomyopathy patients (KS test p=0.04). At a gene level, only one gene, *CPT2*, harbored a 10-fold higher burden of deleterious variants in male compared with female patients (p=0.028), while *MYH6* harbored a 12-fold higher variant burden in female versus male patients albeit this difference was only seen in DCM patients (p=0.024) (**Figure 7g**). There were no striking differences in variant burden by gene group between male and female patients (**Figure 7h**). Overall, these findings suggest that female patients with cardiomyopathy are more likely to harbor variants in constrained coding regions although it is unclear if these differences alone are sufficient to account for sex-related differences in clinical disease severity.

**Figure 7.**
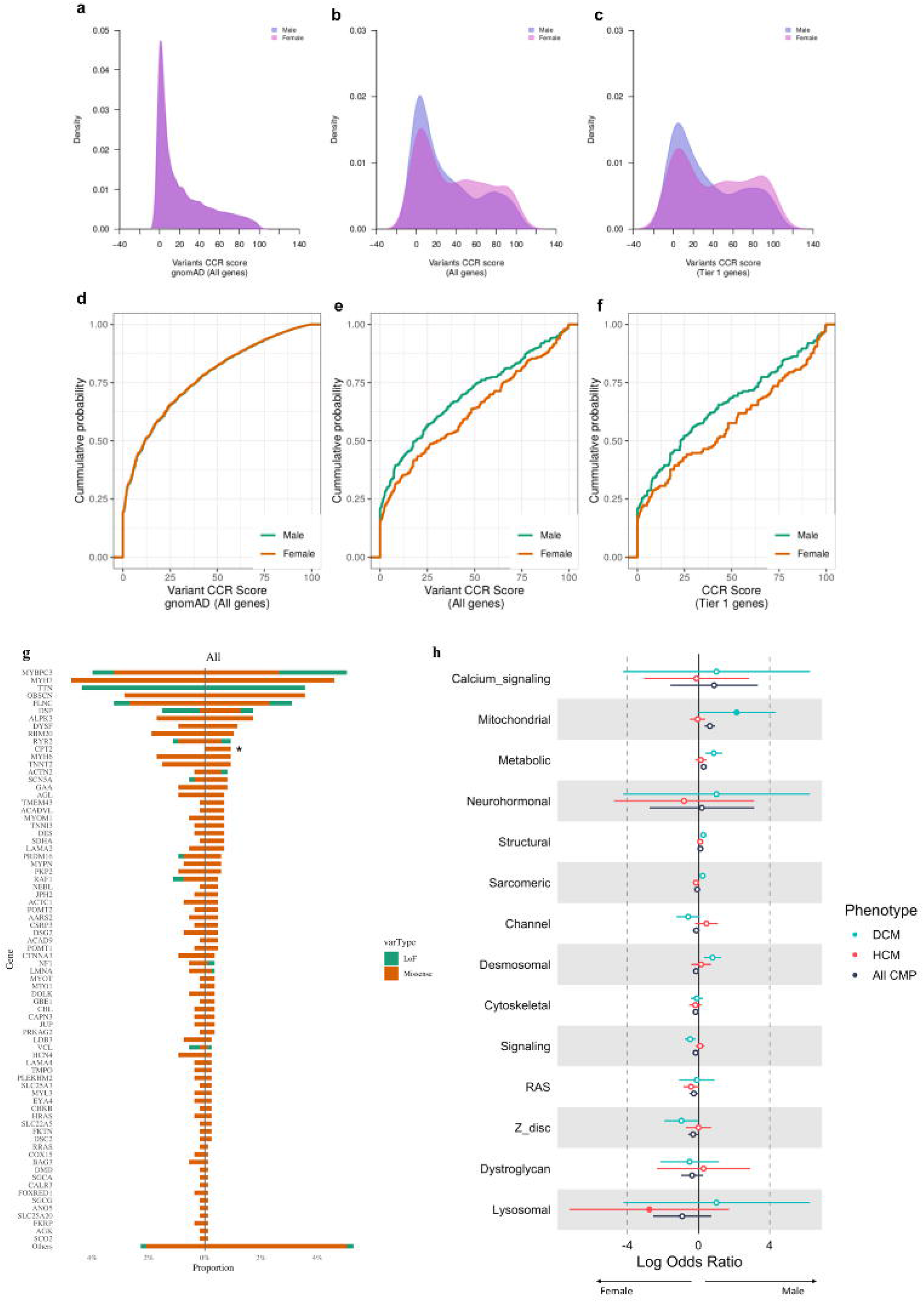
CCR score and affected genes harboring deleterious variants in males and females. (**a-c**) Density plots of CCR scores, and (**d-f**) Empirical cumulative probability distribution of CCR scores of deleterious variants identified in 1397 cardiomyopathy cases and in gnomAD reference controls (n=125,748) stratified by sex. (**d**) There was no difference in variant CCR scores between male and female reference controls in gnomAD. In cardiomyopathy cases, variant CCR scores were higher in female (n=526) compared to male patients (n=871) (**e**) across all cardiomyopathy genes (KS test p=0.002) and (**f**) in the subset of Tier 1 genes (p=0.019). (**g**) Bar plots showing genes harboring rare deleterious variants across all cardiomyopathy patients (871 males, 526 females). *p<0.05 between male and female patients. (**h**) The forest plot shows the log odds ratio comparing proportion of variant carriers between male and female patients for different gene categories across cardiomyopathy subtypes. Except for more male patients harboring more *CPT2* variants (p=0.028), there was no difference in affected genes or gene categories between male and female patients. CCR, constrained coding regions; CMP, cardiomyopathy; gnomAD, Genome Aggregation Database; CMP, cardiomyopathy (black); DCM, dilated cardiomyopathy (blue), HCM, hypertrophic cardiomyopathy (red)

## Discussion

The genetic architecture of cardiomyopathy is heterogeneous, and disease-causing variants have been identified in over 100 genes. By analyzing variants in known cardiomyopathy genes using WGS, we found a higher genetic yield in pediatric compared to adult patients. Importantly, we found age-related differences in affected genes and proteins and in the distribution of variants within constrained coding regions between childhood and adult cardiomyopathy patients that can have implications for using genetic information to guide predictions of age of disease onset and severity.

A striking finding was that variants in childhood cardiomyopathy clustered within highly constrained coding regions of cardiomyopathy genes compared with those identified in adult patients. This was particularly true of sarcomeric gene variants and variants in Tier 1 cardiomyopathy genes. Additionally, deleterious variants in *MYH7* in childhood cases clustered within myosin head and neck regions with very few variants seen in the tail region unlike adult cases. The head and neck regions of MYH7 contain important domains – ATPase site, actin binding site, the converter, and essential light chain, that play a critical role in myocardial contraction and energetics.^36, 37^

To date, truncating variants in the giant sarcomeric gene, titin, located in the A-band region of the gene, have been reported as the most common genetic cause of adult DCM accounting for 20-25% of cases.^33-35, 38, 39^ Our findings confirmed that *TTN* truncating variants were the leading cause of DCM in adult patients but were rare in pediatric patients. Interestingly, *OBSCN* emerged as the second most mutated gene in adult DCM but was rarely mutated in pediatric DCM. Similar to TTN which is a giant protein, OBSCN is a giant protein in the sarcomere that interacts with the z-disk region of TTN. While *OBSCN* missense and frameshift mutations have been shown to co-segregate with HCM, DCM, and LVNC,^40, 41^ our study reveals clustering of *OBSCN* deleterious variants in the protein kinase domains that interact with *CDH2* known to cause ACM.^42^ Taken together, this finding highlights *OBSCN* as an important contributor to adult cardiomyopathy.

Sex-related phenotype differences have been reported in cardiomyopathy with usually a higher prevalence, earlier onset and greater disease severity in male patients.^43-46^ However, we did not detect significant sex-related differences in the frequency and type of affected genes.

Interestingly, overall deleterious variants identified in female patients had higher constraint scores compared with those identified in male patients. This difference was not seen in the reference population. Specifically, deleterious variants in LoF intolerant genes clustered in more constrained coding regions in female patients compared to those identified in male patients.

Taken together, these findings suggest that variants, specially LoF variants in LoF intolerant genes seen in females may be under purifying selection due to greater deleterious effects.

The study has some limitations. The precise age at diagnosis was not known in the adult cases and therefore analysis using age as a linear variable could not be performed. Also, detailed clinical outcomes were not available in the adult cohort, therefore analysis of genotype with outcomes could not be performed. This analysis was limited to coding SNVs and insertion deletions and did not include copy number variants, structural variants or non-coding regulatory variants.

Overall, our study identified deleterious coding variants in *MYH7, MYL3, TNNT2* and *VCL* as more likely to be associated with childhood onset cardiomyopathy, while variants in *TTN* and *OBSCN* were more likely to be associated with adult onset cardiomyopathy. The higher genomic constraints on variants found in pediatric patients and in some female patients suggest that they may be under purifying selection in the general population due to greater deleterious effects.

Overall, our findings suggest that affected gene, variant location within the gene, and variant constraint scores may be useful in predicting early versus late onset cardiomyopathy. This knowledge may inform risk stratification by genotype and help with predictive counseling of affected families.

## Data Availability

Sequencing data is being deposited to the European Genome-Phenome Archive (EGA).

## Acknowledgement

We acknowledge the Labatt Family Heart Centre Biobank at the Hospital for Sick Children for access to DNA samples for whole genome sequencing, and The Centre for Applied Genomics at the Hospital for Sick Children for performing whole genome sequencing. This research was made possible through access to the data and findings generated by the 100,000 Genomes Project. The 100,000 Genomes Project uses data provided by patients and collected by the National Health Service as part of their care and support.

## Consortia

Genomics England Research Consortium

Ambrose, J. C.^1^; Arumugam, P.^1^; Bleda, M.^1^; Boardman-Pretty, F.^1,2^; Boustred, C. R.^1^; Brittain, H.^1^; Caulfield, M. J.^1,2^; Chan, G. C.^1^; Fowler, T.^1^; Giess A.^1^; Hamblin, A.^1^; Henderson, S.^1,2^; Hubbard, T. J. P.^1^; Jackson, R.^1^; Jones, L. J.^1,2^; Kasperaviciute, D.^1,2^; Kayikci, M.^1^; Kousathanas, A.^1^; Lahnstein, L.^1^; Leigh, S. E. A.^1^; Leong, I. U. S.^1^; Lopez, F. J.^1^; Maleady-Crowe, F.^1^; Moutsianas, L.^1,2^; Mueller, M.^1,2^; Murugaesu, N.^1^; Need, A. C.^1,2^; O‘Donovan P.^1^; Odhams, C. A.^1^; Patch, C.^1,2^; Perez-Gil, D.^1^; Pereira, M.B.^1^; Pullinger, J.^1^; Rahim, T.^1^; Rendon, A.^1^; Rogers, T.^1^; Savage, K.^1^; Sawant, K.^1^; Scott, R. H.^1^; Siddiq, A.^1^; Sieghart, A.^1^; Smith, S. C.^1^; Sosinsky, A.^1,2^; Stuckey, A.^1^; Tanguy, M.^1^; Thomas, E. R. A.^1,2^; Thompson, S. R.^1^; Tucci, A.^1,2^; Walsh, E.^1^; Welland, M. J.^1^; Williams, E.^1^; Witkowska, K.^1,2^; Wood, S. M.^1,2^.

^1^. Genomics England, London, UK

^2.^ William Harvey Research Institute, Queen Mary University of London, London, EC1M 6BQ, UK.

